# Reliably quantifying the severity of social symptoms in children with autism using ASDSpeech

**DOI:** 10.1101/2023.10.27.23297600

**Authors:** Marina Eni, Michal Ilan, Analya Michaelovski, Hava M. Golan, Gal Meiri, Idan Menashe, Ilan Dinstein, Yaniv Zigel

## Abstract

Several studies have demonstrated that the severity of social communication problems, a core symptom of Autism Spectrum Disorder (ASD), is correlated with specific speech characteristics of ASD individuals. This suggests that it may be possible to develop speech analysis algorithms that can quantify ASD symptom severity from speech recordings in a direct and objective manner. Here we demonstrate the utility of a new open-source AI algorithm, ASDSpeech, which can analyze speech recordings of ASD children and reliably quantify their social communication difficulties across multiple developmental timepoints. The algorithm was trained and tested on the largest ASD speech dataset available to date, which contained 99,193 vocalizations from 197 ASD children recorded in 258 Autism Diagnostic Observation Schedule, 2^nd^ edition (ADOS-2) assessments. ASDSpeech was trained with acoustic and conversational features extracted from the speech recordings of 136 children, who participated in a single ADOS-2 assessment, and tested with independent recordings of 61 additional children who completed two ADOS-2 assessments, separated by 1–2 years. Estimated total ADOS-2 scores in the test set were significantly correlated with actual scores when examining either the first (*r*(59) = 0.544, *P* < 0.0001) or second (*r*(59) = 0.605, *P* < 0.0001) assessment. Separate estimation of social communication and restricted and repetitive behavior symptoms revealed that ASDSpeech was particularly accurate at estimating social communication symptoms (i.e., ADOS-2 social affect scores). These results demonstrate the potential utility of ASDSpeech for enhancing basic and clinical ASD research as well as clinical management. We openly share both algorithm and speech feature dataset for use and further development by the community.

## Introduction

Autism Spectrum Disorder (ASD) is diagnosed by the presence of social communication difficulties and the existence of Restricted and Repetitive Behaviors (RRBs)^1^. Most ASD children exhibit language delays during early childhood^2^, with 25–30% remaining minimally verbal (i.e., use < 50 words) throughout childhood^3^. However, core ASD symptoms are not necessarily evident in the amount of speech produced by an individual and may instead be evident in the way they speak. Some ASD children exhibit poorer fluency^4^, echolalia (i.e., speech repetition)^5^, mix pronouns^6^, and use atypical articulation and prosody^7,8^ that are apparent in the acoustic features of their vocalizations^9,10^. Studies have reported, for example, that verbal ASD children tend to speak with higher pitch and larger pitch variability than typically developing (TD) children^8,9^. ASD children also exhibit significantly fewer phoneme vocalizations^11^, fewer conversational turns (i.e., reciprocating in a conversation)^11–13^, more non-speech vocalizations^12,14^, more distressed vocalizations (crying, screaming)^15^, and a lower ratio of syllables to vocalizations^16^ than TD children.

Several studies have used automated speech analysis techniques to classify ASD and TD children based on extracted speech features^17–24^. In some studies, diagnostic classification was based on linguistic features such as vocabulary and fluency^24^ while in others it was based on acoustic features such as pitch^18–20,22,23^, jitter^20,23^, shimmer^20,23^, energy^18,19^, Zero-Crossing Rate (ZCR)^18,19^, and Mel-Frequency Cepstral Coefficients (MFCCs)^19^.

Three recent studies have extended this research by training machine and deep learning algorithms to estimate ASD severity according to extracted speech features. In all these studies ground truth was established by clinicians using the Autism Diagnostic Observation Schedule 2^nd^ edition (ADOS-2), a semi-structured assessment where clinicians score the behavior of children during specific tasks/games^25^. The ADOS-2 yields a total severity score as well as separate Social Affect (SA) and Restricted and Repetitive Behaviors (RRB) scores that quantify social difficulties and RRB symptoms, respectively. In the first study, the authors extracted vocalization rates and durations from speech recordings of 33 ASD children during an ADOS-2 assessment and reported that a trained synthetic random forest model was able to accurately estimate their ADOS-2 Social Affect (SA) scores^26^. Another study extracted hundreds of conversational, acoustic, and lexical speech features from speech recordings of 88 adolescents and adults with ASD during an ADOS assessment (1^st^ edition) and reported that a trained Deep Neural Network (DNN) was able to accurately estimate scores of four specific ADOS items that quantify the ability to maintain a mature social conversation^27^. Finally, in the third study, from our group, we extracted acoustic features such as pitch and energy, and conversational features such as turn-taking and speech rate from speech recordings of 72 children (56 with ASD) during an ADOS-2 assessment^28^. We demonstrated that a trained Convolutional Neural Network (CNN) model was able to accurately estimate total ADOS-2 scores across multiple train-test subsamples.

While these results are encouraging, algorithms developed so far were trained and tested with relatively small ASD samples that are not likely to represent the large heterogeneity of speech styles and characteristics in the broad ASD population^29^. Moreover, previous studies examined only a single timepoint of data from each participant, thereby limiting the ability to assess the reliability of algorithms to assess ASD symptom severity at different developmental timepoints. Previous studies also did not compare the ability of deep learning models to successfully estimate the severity of social ASD symptoms versus RRB symptoms. Most importantly, previous studies did not share their algorithms and data in a transparent manner that would enable re-production of results and further development of algorithms by the research community.

To address these limitations, we created the largest speech recording dataset available to date, which contained 99,193 vocalizations from 197 ASD children recorded in 258 ADOS-2 assessments, with 61 of the children participating in two ADOS-2 assessments that were separated by 1-2 years. This comprehensive dataset enabled us to train and test the ASDSpeech algorithm on different subsets of children and compare its accuracy across two developmental timepoints as well as sex and age sub-groups. In addition, we also examined the ability to estimate ADOS-2 SA versus RRB scores (i.e., social difficulties versus RRB symptoms). We intentionally used raw ADOS-2 scores, which have a considerably wider range than ADOS-2 calibrated severity scores ^30,31^, thereby increasing the potential sensitivity of the algorithm. Finally, we openly share the algorithm and speech feature dataset to promote transparency and enable further use and development by the research community.

## Methods

### Participants and setting

We analyzed data collected at the Azrieli National Centre for Autism and Neurodevelopment Research (ANCAN), a collaboration between Ben-Gurion University of the Negev (BGU) and eight partner clinical centers where ASD is diagnosed throughout Israel. ANCAN manages the national autism database of Israel with data from > 3000 children in 2023 and growing^32,33^. All recordings used in the current study were performed in a single ANCAN assessment room located at Soroka University Medical Center (SUMC), the largest partner clinical site. A total of 197 children (1–7-years-old) who completed at least one ADOS-2 assessment between 2015 and 2021 and received an ASD diagnosis were included in this study (Table 1). Of the participating children, 136 completed a single ADOS-2 assessment and 61 completed two ADOS-2 assessments at two timepoints separated by 10–29 months, yielding 258 ADOS-2 assessments in total. All ADOS-2 assessments were performed by a clinician with research reliability. In addition, all participating children had ASD diagnoses that were confirmed by both a developmental psychologist and either a child psychiatrist or a pediatric neurologist, according to Diagnostic and Statistical Manual of Mental Disorders, Fifth Edition (DSM-5) criteria^34^. Informed consent was obtained from all parents, and the study received approval from the SUMC Helsinki committee.

**Table 1.**
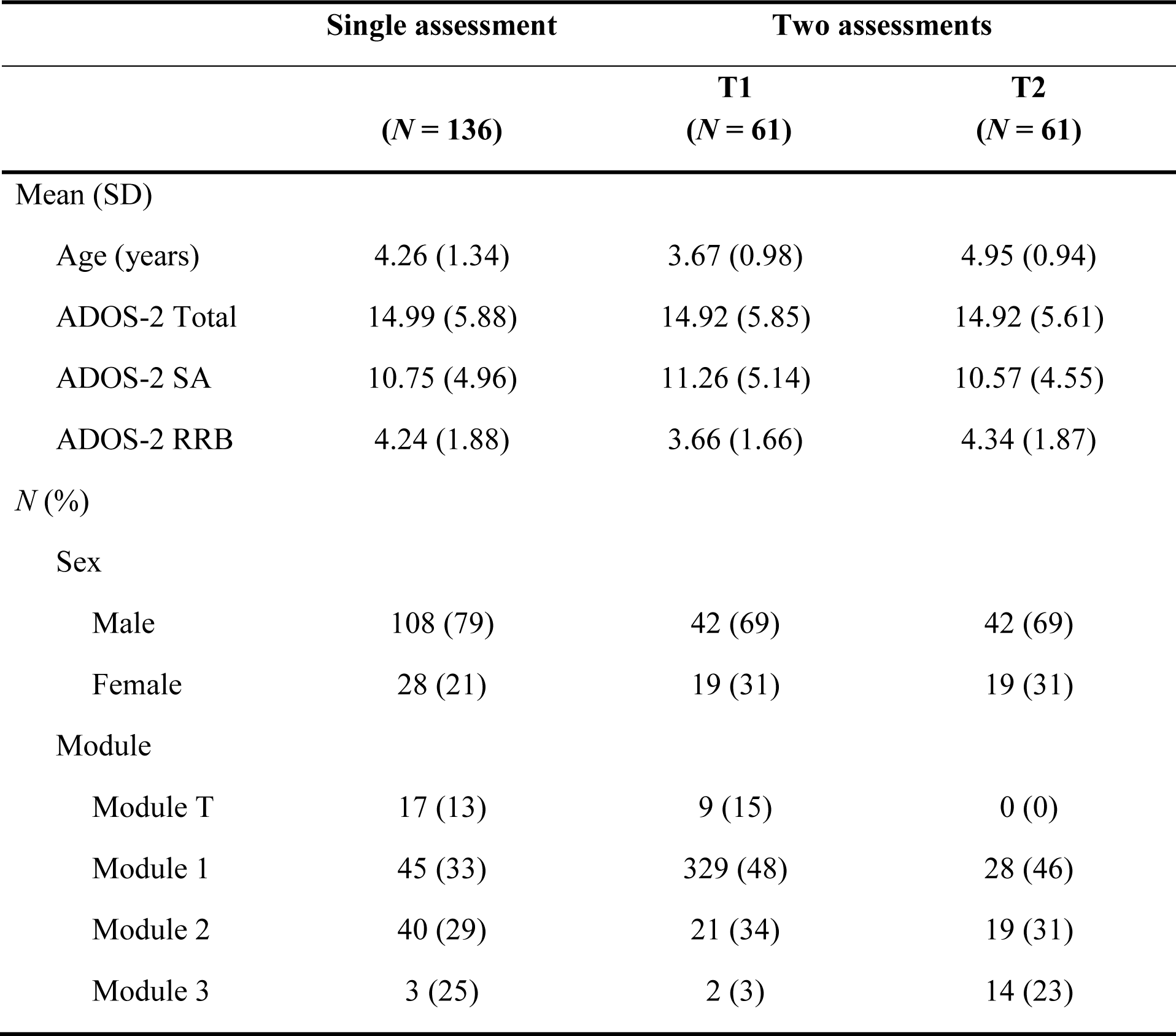
Participating children’s characteristics.

### ADOS-2 assessments

ADOS-2 is a semi-structured behavioral assessment where a clinician administers specific tasks, observes the behavior of the child, and scores their behavior^35^. The total ADOS-2 score (range: 0–30) is the sum of the Social Affect (SA, range: 0–22) and Restricted and Repetitive Behavior (RRB, range: 0–8) scores, with higher scores indicating more severe symptoms.

### Recording setup

All ADOS-2 recordings were performed using a single microphone (CHM99, AKG, Vienna) located on a wall, ∼1–2m from the child, and connected to a sound card (US-16×08, TASCAM, California). Each ADOS-2 session lasted ∼40-minutes (40.75 ± 11.95 min) and was recorded at a sampling rate of 44.1 kHz, 16 bits/sample (down-sampled to 16 kHz).

### Detection of child vocalizations

We manually labeled segments with child vocalizations in each of the audio recordings. These segments included speech, laughing, moaning, crying, and screaming. The child segments often contained multiple vocalizations (e.g., multiple utterances) separated by silence. We separated each segment into multiple vocalizations using energy thresholds of 2.79dB and 0.4dB above the background noise to define the beginning and end of each vocalization, respectively^28^ (Supplementary Figure S1). Vocalizations that were shorter than 110ms were excluded from further analysis (too short to contain an utterance).

### Features

We extracted 49 speech features from the child vocalizations that were categorized into nine groups: pitch, formants, jitter, voicing, energy, Zero-Crossing Rate (ZCR), spectral slope, duration, and quantity/number of vocalizations. All features, except duration and quantity, were first extracted in 40ms windows (window overlap of 75%), resulting in a vector of feature values per vocalization. The minimum, the maximum, and the mean pitch of the voiced vocalizations (across windows) were computed, deriving one value for each vocalization. We then selected a group of 10 consecutive vocalizations and computed the mean and variance across vocalizations for relevant features (Supplementary Table S1). We also computed the mean duration of vocalizations and the overall number of vocalizations in the recording. Taken together, these steps yielded a vector with 49 values corresponding to the 49 features per 10 vocalizations. We performed this procedure 100 times, selecting random groups of ten consecutive vocalizations from the recording. Combining these 100 samples yielded a features matrix of 100×49 per child (Supplementary Figure S2), with the last column (quantity of vocalizations) containing the same value across all rows. Features included:

**Frequency related features:**

- *Pitch (F0):* Vocal cords vibration frequency (the fundamental frequency) that exists only in voiced speech (e.g., vowels). Voiced Vocalization (VV) was defined as a vocalization where most of its frames (≥ 60%)^10^ were voiced (voicing threshold 0.45).
- *Formants:* The resonant frequencies of the vocal tract that shape vowel sounds^36^. The first two formants (F1 and F2) relate to tongue position (vertical and horizontal) and influence vowel quality. Their bandwidths affect the clarity of speech.
- *Jitter:* Variation across adjacent pitch values representing frequency instability^37^.
- *Voicing*: Pitch peak amplitude as determined by the autocorrelation function.

Pitch and formants were calculated using the PRAAT software^38^, with a pitch range set to 60–1600 Hz (a wide range to increase sensitivity to atypical vocal characteristics).

**Energy/amplitude related features:**

- *Energy:* The energy ratio between each child’s vocalization and the background noise. The background noise energy was calculated from the energy values extracted from the lowest 5% of the recording’s frames.

**Spectral features:**

- *Zero-Crossing Rate (ZCR):* The number of zero-crossings apparent in audio segments with child vocalizations^39^.
- *Spectral slope:* The slope of the linear regression on the logarithmic power spectrum within the frequency bands of 20–500 Hz (lower band) and 500–1500 Hz (higher band)^40,41^.

**Conversational features:**

- *Duration:* Child’s mean vocalization length.
- *Quantity*: The total number of vocalizations.

All features, except for Pitch and Formants, were extracted with custom-written code in Matlab (Mathworks, Inc.).

### Training and testing ASDSpeech

Training was performed with data from the 136 children who completed a single ADOS-2 session only. Feature matrices were used to train two deep learning models with an identical CNN architecture (Supplementary Figure S3). The first model estimated ADOS-2 SA scores and the second estimated ADOS-2 RRB scores. Training was based on minimizing the Mean Squares Error (MSE) of a regression analysis between estimated and actual scores, using the RMSprop (Root Mean Square Propagation) as the optimization algorithm^42^. The training process was preformed 25 times, creating 25 different SA and 25 RRB models that were trained with different combinations of training data sub-samples and learning parameters. We considered this analogous to having 25 clinicians, each with a different learning style and different clinical experience. First, we performed the feature extraction procedure described above 5 times for each child. Since feature extraction included a random selection of consecutive vocalizations, this resulted in 5 different sub-samples of the data. When training each model (separately for SA and RRB) we split the training data into a training-set (80%) and validation set (20%) and applied a random search algorithm to optimize the following learning parameters: batch size, number of epochs, and learning rate, while applying early stopping of patience after 20 epochs to reduce overfitting^43^. Optimal learning hyper-parameters were selected based on the highest concordance correlation coefficient^44^, between estimated and actual ADOS-2 scores in the training and validation sets respectively. This procedure was performed 5 times using different selections of validation data (i.e., 5-fold cross validation), yielding 5 models with different learning parameters per data sub-sample and 25 models in total for SA and RRB scores separately.

Testing was performed with an entirely independent dataset of 61 ASD children who completed two ADOS-2 assessments. For each of these children we estimated a separate SA and RRB score from each of the 25 models described above and then computed their mean, yielding a single SA and RRB score per child. This is analogous to a clinical consensus across the 25 models. Accuracy of ASDSpeech estimation was measured using Pearson correlation and NRMSE (RMSE / (*y*_max_ - *y*_min_), where *y* is the actual ADOS-2 score), which were calculated between the estimated and actual ADOS-2 scores in the testing dataset, separately for the first and second ADOS-2 assessments (i.e., T1 and T2).

### Hardware

All model training, optimization, and training were performed using custom-written code in Python 3.9.13 using a Keras API 2.6.0 with TensorFlow (version 2.6.0) backend. The training was conducted on an IntelI XI(R) Gold 6140 CPU @ 2.30GHz and NVIDIA GPU Tesla T4.

### Statistical Analysis

All statistical analyses were conducted using custom-written code in Python. Associations between speech features and ADOS-2 scores were assessed using Pearson correlations. To evaluate their statistical significance, we performed a random permutation test. In this test, we randomly shuffled the actual ADOS-2 scores across children and calculated the correlation between each feature and the shuffled scores. This randomization procedure was performed 1,000 times, generating a null distribution of random correlation values as computed from the original data that is not necessarily normally distributed as assumed by parametric statistical tests. For a correlation between a speech feature and ADOS-2 score to be considered significant, the actual correlation value had to be higher than the 97.5 percentile of the null distribution. We used an equivalent analysis to assess the statistical significance of correlations between actual and ASDSpeech estimated ADOS-2 scores. We also performed a similar analysis with NRMSE values, where we assessed whether the actual NRMSE value was smaller than the 2.5 percentile of the null distribution. This statistical test, therefore, assessed whether correlation values were higher than expected by chance and NRMSE values were lower than expected by chance.

### Data sharing

The ASDSpeech algorithm source-code and associated dataset are available at https://github.com/Dinstein-Lab/ASDSpeech.

## Results

Using the data from the 136 ASD children in the training dataset, we examined the relationships between each of the 49 features and ASD symptom severity as defined clinically by the children’s ADOS-2 scores. Thirty-one features exhibited significant Pearson correlation coefficients with total ADOS-2 scores (i.e., sum of SA and RRB scores), 31 with ADOS-2 SA scores, and 28 with ADOS-2 RRB scores (Figure 1). While some features, such as the number of vocalizations, exhibited a stronger correlation with SA than RRB score, others, such as mean jitter, exhibited the opposite (Supplementary Figure S4). Hence, different features seem to carry distinct information regarding each of the two core ASD symptoms, demonstrating the potential opportunity for a deep learning algorithm to learn relevant associations.

**Figure 1.**
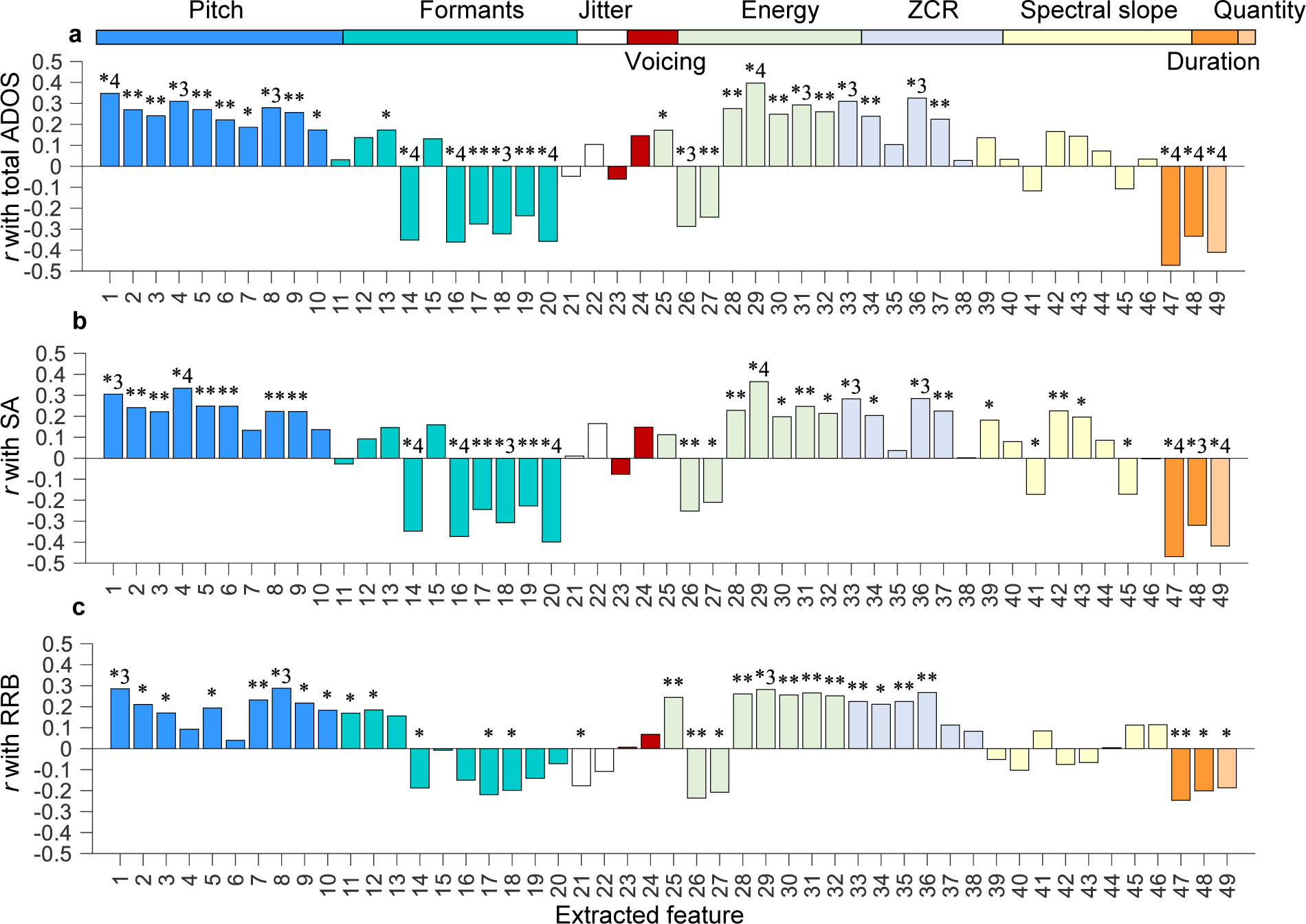
Pearson correlation coefficients between each of the extracted features and ADOS-2 scores from the 136 children in the training dataset. Correlation coefficients are presented for total ADOS-2 scores (a), ADOS-2 SA scores (b), and ADOS-2 RRB scores (c). Each color represents a different group of features. Asterisks: significant Pearson correlation (∗ < 0.05, ∗∗ ≤ 0.01, ∗3 ≤ 0.001, ∗4 ≤ 0.0001).

### Longitudinal stability of ADOS-2 scores

The 61 ASD children in the test dataset exhibited similar ADOS-2 scores across their two assessments, which were separated by 1-2 years, indicating overall stability in severity over time. Significant correlations were apparent across first and second assessments for ADOS-2 total (*r*(59) = 0.743, *P* < 0.001), ADOS-2 SA (*r*(59) = 0.666, *P* < 0.001), and ADOS-2 RRB (*r*(59) = 0.5, *P* < 0.001) scores (Figure 2).

**Figure 2.**
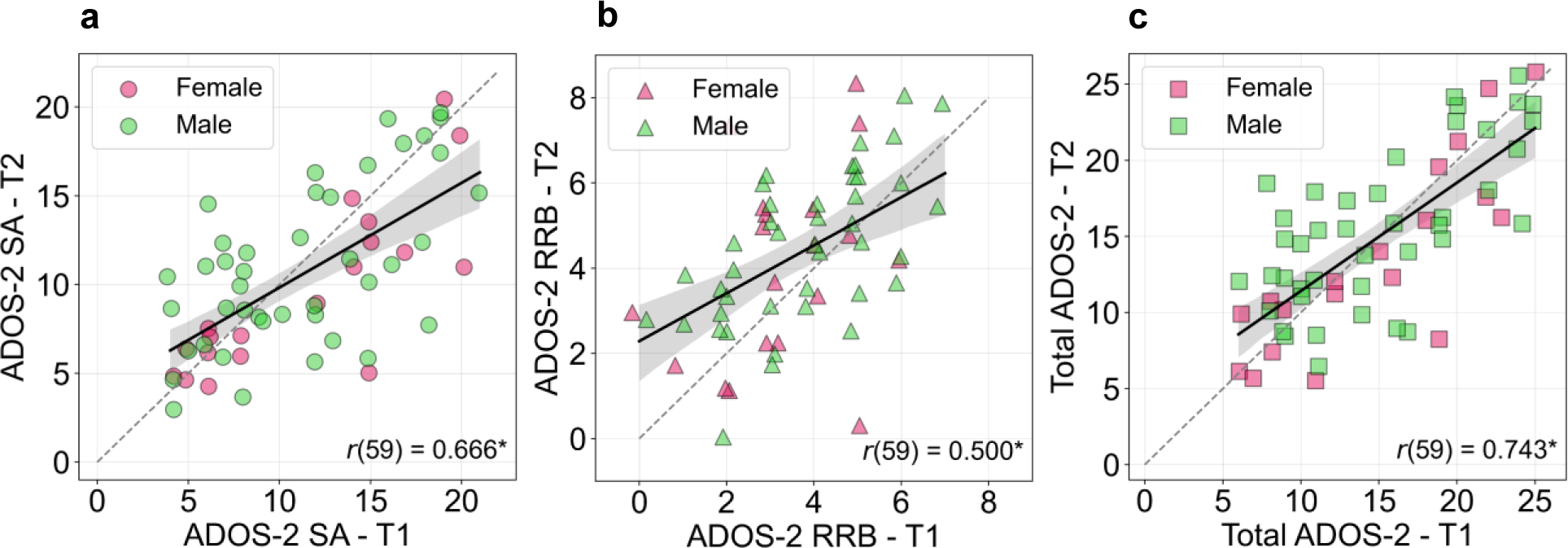
Scatter plots demonstrating overall stability in ADOS-2 scores across first and second assessments (T1 and T2). (**a**) ADOS-2 SA scores. (**b**) ADOS-2 RRB scores. (**c**) Total ADOS-2 scores (sum of SA and RRB scores). **Asterisk**: statistical significance of the Pearson correlation coefficient (*P* < 0.0001). **Shaded areas**: 95% confidence intervals. Children located below the diagonal (dashed line) exhibited lower ASD severity at T2 (improvement), while children above the diagonal exhibited the opposite.

### Training and testing the ASDSpeech algorithm

We trained the ASDSpeech algorithm with data from 136 ASD children in the training dataset. The algorithm included two separate CNN models that were trained to estimate ADOS-2 SA and RRB scores independently, given that different speech features were associated with each symptom domain. The accuracy of the algorithm was tested with data from two independent ADOS-2 recordings of the 61 children in the testing dataset where ASDSpeech estimated the SA, RRB, and total ADOS-2 (sum of SA and RRB) scores of each child per recording (Figure 3).

**Figure 3.**
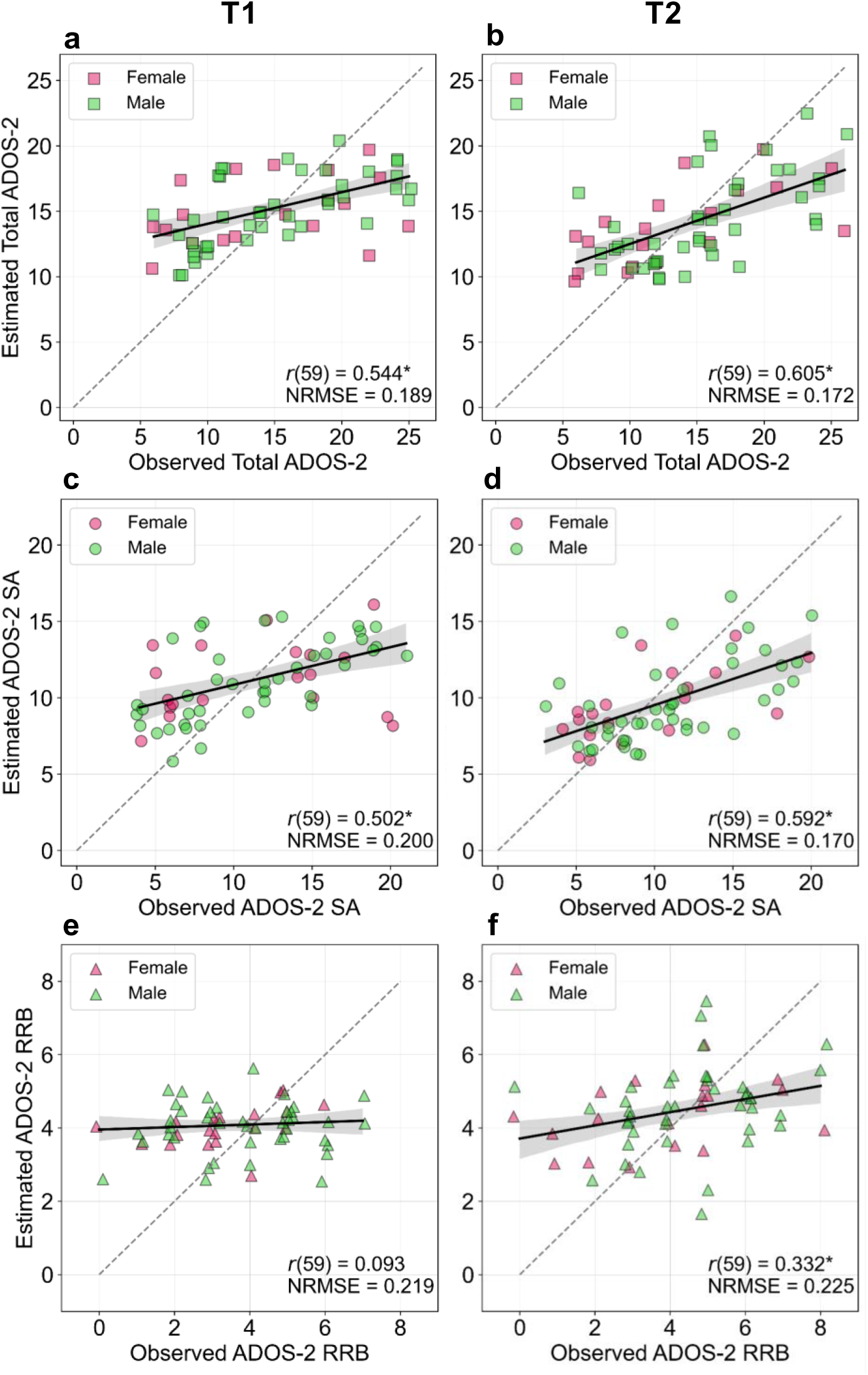
Accuracy of ASDSpeech. Scatter plots demonstrating the fit between estimated and actual scores for the children at T1 (left column) and T2 (right column). **(a-b)** Total ADOS-2 scores (sum of SA and RRB scores). (**c-d**) ADOS-2 SA scores. (**e-f**) ADOS-2 RRB scores. Pearson correlation coefficients and NRMSE values are noted in each panel. **Solid line:** Linear fit. **Dashed line:** diagonal (unity line). **Asterisks:** statistical significance as determined by randomization test (*P* < 0.05).

Estimated total ADOS-2 scores were significantly correlated with actual scores at T1 (*r*(59) = 0.544, *P* < 0.0001) and T2 (*r*(59) = 0.605, *P <* 0.0001). Similarly, estimated ADOS-2 SA scores were significantly correlated with actual scores at T1 (*r*(59) = 0.502, *P* < 0.0001) and T2 (*r*(59) = 0.592, *P* < 0.0001). In contrast, estimated ADOS-2 RRB scores were not significantly correlated with actual RRB scores at T1 (*r*(59) = 0.093, *P* = 0.474), exhibiting significant correlations only at T2 (*r*(59) = 0.332 *P* = 0.009) with a relatively weaker effect size.

Normalized Root Mean Squared Error (NRMSE) between estimated and actual total ADOS-2 scores was significantly smaller than expected by chance when computed at T1 (NRMSE = 0.189, *P* < 0.0001) and T2 (NRMSE = 0.172, *P =* 0.0001). Similarly, NRMSE between estimated and actual ADOS-2 SA scores was significantly smaller than expected by chance when computed at T1 (NRMSE = 0.200, *P* < 0.0001) and T2 (NRMSE = 0.170, *P* < 0.0001). In contrast, NRMSE between estimated and actual ADOS-2 RRB scores was not significantly smaller than expected by chance at T1 (NRMSE = 0.219, *P* = 0.460), exhibiting significant results only at T2 (NRMSE = 0.225, *P* = 0.006).

The statistical significance of the NRMSE results was determined with a randomization analysis where we randomly shuffled ADOS-2 scores across children before computing NRMSE values. We computed 1,000 random permutations to generate a null NRMSE distribution and assessed statistical significance by determining whether the actual NRMSE value was smaller than the 2.5 percentile of the null distribution (see Methods).

### Differences across age and sex subgroups

Next, we examined whether ASDSpeech accuracy differed across age and sex subgroups (Figure 4). Estimated total ADOS-2 scores were significantly correlated with actual scores when examining children above the median age at T1 (*r*(28) = 0.604, *P* = 0.0004) or T2 (*r*(25) = 0.612, *P* = 0.0007) and children below the median age at T1 (*r*(29) = 0.485, *P* = 0.006) or T2 (*r*(32) = 0.657, *P* < 0.0001). There were no significant differences in the algorithm’s accuracy between younger and older children at T1 (*P* = 0.540) or T2 (*P* = 0.780) as tested with a randomization analysis. Similarly, estimated total ADOS-2 scores were significantly correlated with actual scores when examining males at T1: (*r*(40) = 0.631, *P* < 0.0001) or T2 (*r*(40) = 0.601, *P* < 0.0001). Estimated ADOS-2 scores were also significantly correlated with actual scores when examining females at T2 (*r* (17) = 0.627, *P* = 0.004), but the correlation did not reach statistical significance at T1 (*r* (17) = 0.363, *P* = 0.127). Nevertheless, there were no significant differences in the algorithm’s accuracy between males and females at T1 (*P* = 0.198) or T2 (*P* = 0.930) as tested with a randomization analysis.

**Figure 4.**
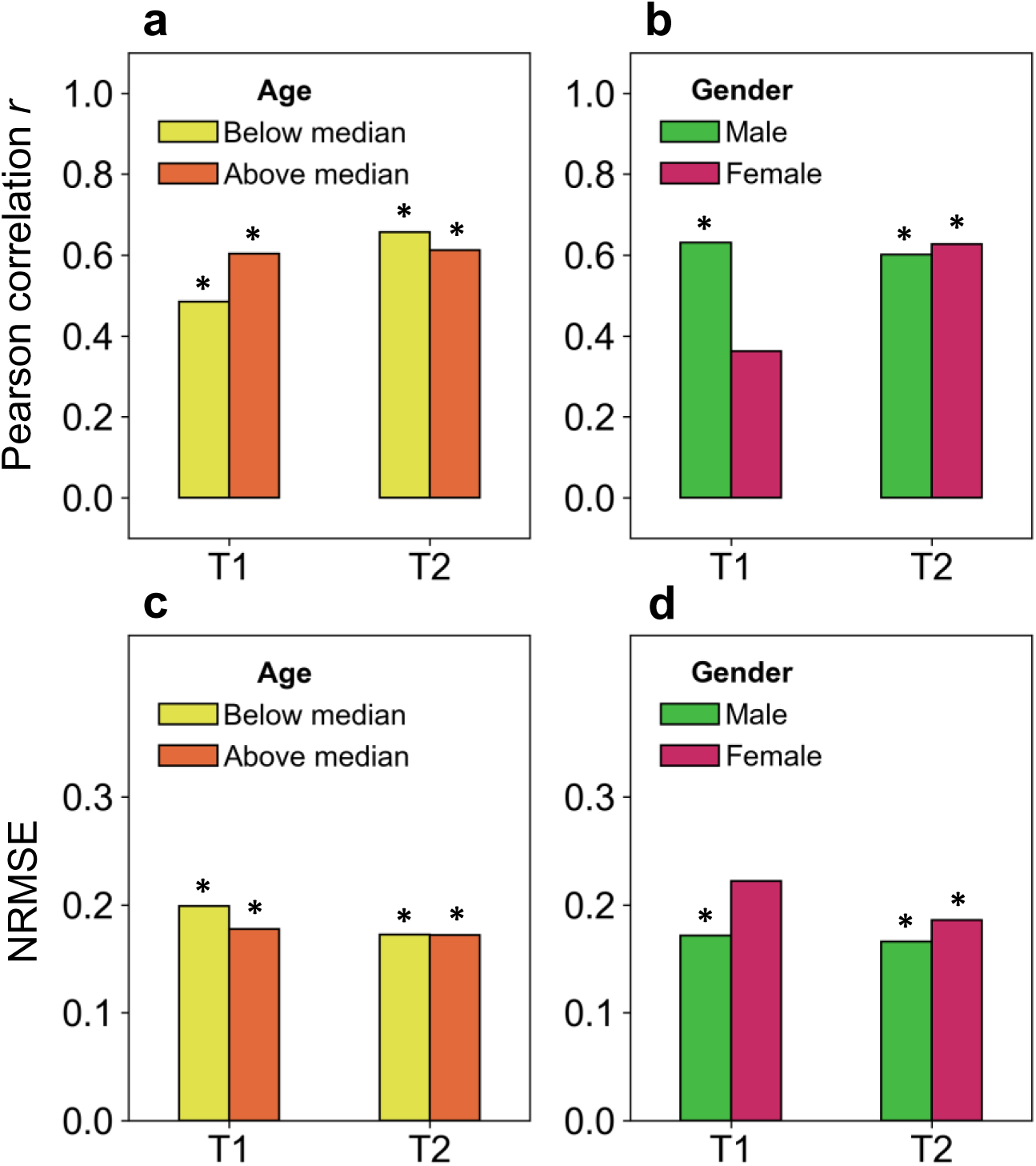
ASDSpeech accuracy as a function of sex and age at T1 and T2. (**a**,**b**) Pearson correlation values (**c,d**) Normalized Root Mean Squared Error (NRMSE) values. (**a**, **c**) comparison between younger and older children (median split according to age at each timepoint). (**b**, **d**) comparison between males and females. **Asterisks:** statistical significance as determined by randomization test (*P* < 0.05).

Comparison of NRMSE across subgroups showed similar results. NRMSE between the estimated and actual ADOS-2 scores was significantly smaller than expected by chance when examining younger children at T1 (NRMSE = 0.199, *P* = 0.008) or T2 (NRMSE = 0.173, *P* < 0.0001) as well as older children at T1 (NRMSE = 0.178, *P* < 0.0001) or T2 (NRMSE = 0.172, *P* < 0.0001). There were no significant differences in the algorithm’s accuracy between younger and older children at T1 (*P* = 0.434) or T2 (*P* = 0.992). NRMSE were also significantly smaller than expected by chance when examining males at T1 (NRMSE = 0.172, *P* < 0.0001) or T2 (NRMSE = 0.166, *P* < 0.0001). For females this was the case only at T2 (NRMSE = 0.186, *P* = 0.006) and not at T1 (NRMSE = 0.222, *P* = 0.140). Nevertheless, there were no significant differences in the algorithm’s accuracy between males and females at T1 (*P* = 0.094) or T2 (*P* = 0.588) as tested with a randomization test.

## Discussion

Our results demonstrate the ability of ASDSpeech to quantify the severity of social symptoms in ASD children from recordings of their speech during ADOS-2 assessments. The algorithm, trained with recordings from 136 ASD children, was able to accurately estimate total ADOS-2 and ADOS-2 SA scores in an entirely independent sample of 61 ASD children, who were recorded at two different developmental timepoints separated by 1-2 years (Figure 3). It is remarkable that ASDSpeech was able to achieve this despite the large heterogeneity in language fluency and speech articulation abilities apparent across ASD children^45^ as well as the large developmental changes that take place in speech abilities during the examined period of early childhood^46^. Moreover, the robust accuracy of ASDSpeech in estimating ADOS-2 SA scores is remarkable given that the social difficulties assessed during the ADOS-2 assessment manifest themselves in behaviors that have little to do with speech including difficulties with eye contact, imitation, joint attention, and other social behaviors^3,47^. This suggests that combining ASDSpeech with analysis of eye tracking^48–50^, facial expressions^51^, and body movement^52^ data from the same children will enable even higher accuracy and reliability in estimating ASD symptoms.

Separate estimation of social and RRB symptoms demonstrated that ASDSpeech was considerably more accurate at estimating social ASD symptoms captured by the ADOS-2 SA scores in contrast to the RRB symptoms captured by the ADOS-2 RRB scores (Figure 3). Note that accurate estimation of total ADOS-2 scores (Figure 3) was likely based on the accurate estimation of SA scores that account for two-thirds of the total scores. We believe there may be several reasons for the more accurate estimation of SA scores. First, the limited range of the ADOS-2 RRB scale (0–8) relative to the SA scale (0–22) may make it difficult for the algorithm to identify differences across children. Indeed, a recent study reported that the limited number of items on the RRB scale resulted in poor scale reliability across participants^53^. Second, the selected speech features in the current study exhibited weaker correlations with RRB than SA scores (Figure 1). Extraction of additional speech features, such as phrase or intonation repetitions (indicative of echolalia) may improve the accuracy of ADOS-2 RRB score estimates. Regardless, our results motivate separate modeling of social and RRB symptom domains as each of them is likely associated with distinct features of speech.

In the current study we estimated raw ADOS-2 scores rather than calibrated severity scores (CSS), which were developed to standardize ASD symptom severity measurements across different ages and language abilities^30,31^. While ADOS-2 CSS are important for longitudinal assessments of coarse changes in severity^54,55^, their restricted scoring range (children with ASD receive scores of 4-10) limits the sensitivity of deep learning algorithms in identifying differences across children. By demonstrating that ASDSpeech achieves robust accuracy in estimating raw ADOS-2 SA scores across different age groups and developmental timepoints we show that severity estimations are independent of these factors, thereby justifying the use of raw scores.

### Diagnostic classification with speech analysis algorithms

A variety of previous studies have reported that individuals with ASD, on average, speak differently than TD individuals^4,8–16^. According to these studies, ASD individuals exhibit atypical speech characteristics, including significantly fewer phonemes per utterance^11^, fewer conversational turns^13^, higher pitch^9,19^, and larger pitch range and variability^8,9^ than TD children. Differences in these and other speech characteristics have enabled the development of machine and deep learning classification algorithms that can identify ASD and TD individuals with reported accuracy rates of 75-98%^17–23^.

However, these relatively high classification accuracies are likely to be inflated due to the small sample size of most studies (<40 ASD participants) that are not likely to capture the true heterogeneity of ASD symptoms or speech styles/characteristics of the broad ASD population. Indeed, even “gold standard” clinical tests such as the ADOS-2, exhibit ∼80% accuracy in identifying children who will eventually receive an ASD diagnosis^56^. This is because establishing an ASD diagnosis requires clinicians to incorporate additional information from parent interviews and other clinical assessments^57^. Clinicians also report high diagnosis certainty in only ∼70% of ASD children because the presentation of ASD symptoms is equivocal in ∼30% of cases^58^. These studies suggest an expected upper limit of 70–80% accuracy when attempting to identify ASD using digital phenotyping techniques such as speech analysis. Nevertheless, it is highly encouraging that speech features contain information enabling the separation of ASD and TD children.

### Quantifying ASD severity with speech analysis algorithms

A more complex task is to develop machine and deep learning algorithms that can quantify the severity core ASD symptoms. Results presented in the current and previous study from our lab^28^ demonstrated that multiple speech features were significantly correlated with SA and/or RRB ADOS-2 scores (Figure 1), suggesting that distinct combinations of speech features are associated with each of the two core ASD symptoms.

Three recent studies have attempted to use these relationships to predict ADOS-2 scores by analyzing speech recordings of ASD individuals^26–28^. The first trained a synthetic random forest model to estimate ADOS-2 SA scores according to vocalization rate and turn-taking features extracted from ADOS-2 recordings of 33 ASD children. The algorithm was able to predict ADOS-2 SA scores that were significantly correlated with actual scores (*r* = 0.634). The second study utilized a DNN model to estimate four ADOS (first edition) item scores using hundreds of conversational and acoustic features extracted from speech recordings of 88 high-functioning ASD adolescents/adults during an ADOS assessment^27^. This algorithm was able to estimate scores that exhibited significant Spearman correlations with the actual scores (*ρ* = 0.519–0.645). Finally, in a previous study from our lab^28^, we demonstrated that a CNN model was able to estimate ADOS-2 total scores that were significantly correlated with actual scores (*r* = 0.718) when using 60 conversational and acoustic features extracted from speech recordings of 72 children (56 of them with ASD) during ADOS-2 assessment.

The current study extends previous work in several critical ways. First, we utilized a considerably larger dataset (258 ADOS-2 recordings) that was at least three times larger than the ones used to date. This was important for training ASDSpeech with speech recordings form a large cohort with heterogeneous language abilities. Second, the 61 ASD children in our testing dataset were recorded twice during two ADOS-2 assessments separated by 1–2 years. This enabled us to test the robustness of ASDSpeech across two developmental timepoints. Third, we trained ASDSpeech to estimate ADOS-2 SA and ADOS-2 RRB scores using separate CNN models. The results demonstrated that this separation was critical with accurate performance apparent primarily for the ADOS-2 SA scores. Fourth, the large sample size enabled us to demonstrate that ASDSpeech accuracy was similar across age and sex subgroups. Fifth, the recordings utilized in the current study were performed over a 6-year period in a busy public healthcare medical center that services a population of ∼1 million people. Recordings were performed with a wall mounted microphone (see Methods) in “real world” noisy conditions (e.g., announcement system in the hallway). This demonstrates the robustness of ASDSpeech to variable recording conditions.

ASDSpeech achieved similar accuracy to that reported in previous studies. The important advance in the current study is in demonstrating that this accuracy is robust to age and developmental stage of the examined children when examining a large heterogeneous population within an active clinical setting. Most importantly, we openly share ASDSpeech and its associated dataset with the research community.

### Limitations

The current study had several limitations. First, we did not examine the language content of the recordings, which is likely to improve the estimation of ASD symptom severity^4,24^. Second, we did not identify echolalia, crying, or shouting events that are likely to be informative of RRB symptoms. Indeed, our weaker results estimating RRB scores suggest that different speech features are necessary for estimating severity in this domain. Third, we did not apply any noise reduction or multi-speaker analysis techniques to improve the quality of the analyzed vocal segments. Finally, our sample had a 4:1 male to female ratio, which is equivalent to the sex ratio in the national ASD population of Israel^59^. Hence, higher ASDSpeech accuracy for males at T1 may be due to the larger number of males in the training and testing datasets. This could be rectified by future studies.

### Conclusions

This study adds to accumulating evidence demonstrating that speech recordings contain reliable information about the social symptom severity of ASD children. We demonstrate the ability of the ASDSpeech algorithm to quantify these symptoms in a robust manner across two developmental timepoints with recordings that were performed within a busy community healthcare center. We openly share the algorithm and its associated dataset for further use, testing, and development by the research community and are confident that future versions of the algorithm will achieve even higher and more robust accuracy rates, yielding a transformative new tool for clinical and basic ASD research.

## Data Availability

The data generated in the current study is not publicly accessible due to ethical constraints

## Acknowledgments

This study was supported by the Israeli Science Foundation (Grant no. 1150/20) and the Israel Ministry of Science & Technology (Grant no. 3-17422).

## Author contributions

M.E. collected the data, performed the experiments, built the models, analyzed the data, and wrote the paper. I.D., and Y.Z. designed the study, guided data collection and analysis, and wrote the paper. M.I., A.M., H.M.G., G.M., and I.M. contributed to participant recruitment, behavioral assessments, data collection, and interpretation of the findings. All authors approved the final manuscript.

## Additional information

### Competing interests

The authors declare no competing interests.

## Notes

### Competing Interest Statement

The authors have declared no competing interest.

### Funding Statement

This study was funded by Israeli Science Foundation (Grant no. 1150/20) and Israel Ministry of Science & Technology (Grant no. 3-17422)

### Author Declarations

Ethics committee/IRB of Soroka Medical Centre Research Ethics Board waived ethical approval for this work

### Summary of Updates

updated supplementary file is uploaded.

