## Supplementary material for "Reliably quantifying the severity of social symptoms in children with autism using ASDSpeech"

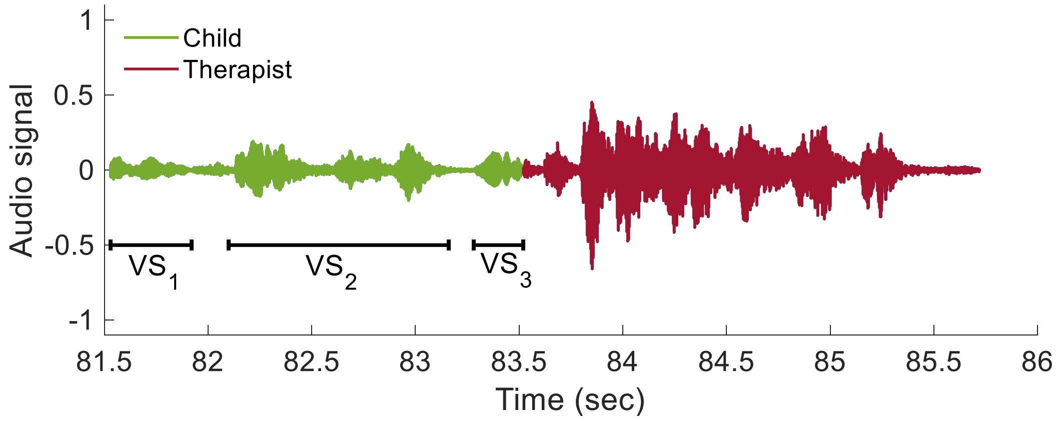


**Supplementary Figure S1. Example of manual segmentation and vocalizations detected in a child's speech segment.** The example includes two manually labeled segments with the first belonging to a child (green, first 4s) and the second to the clinician (red). The child segment was split into three vocalizations according to energy thresholds: $VS_1$, $VS_2$, and $VS_3$.

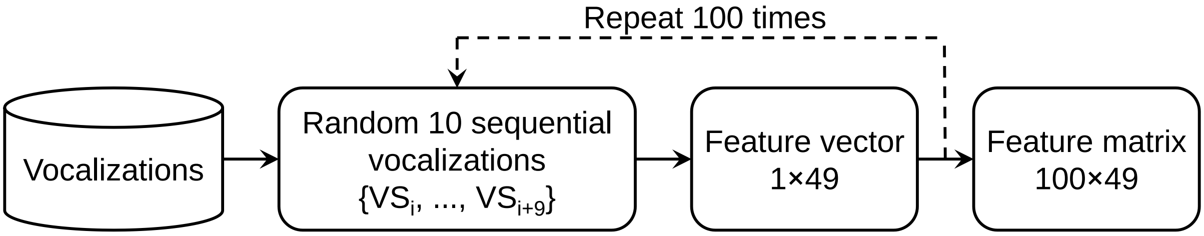


**Supplementary Figure S2: Creating the feature matrix used as input for ASDSpeech.** Groups of 10 consecutive vocalizations were selected and features were extracted across the 10 vocalizations. $VS_i$ is the i$^{th}$ vocalization of group. This process was performed 100 times with different groups of 10 vocalizations to create the input matrix.

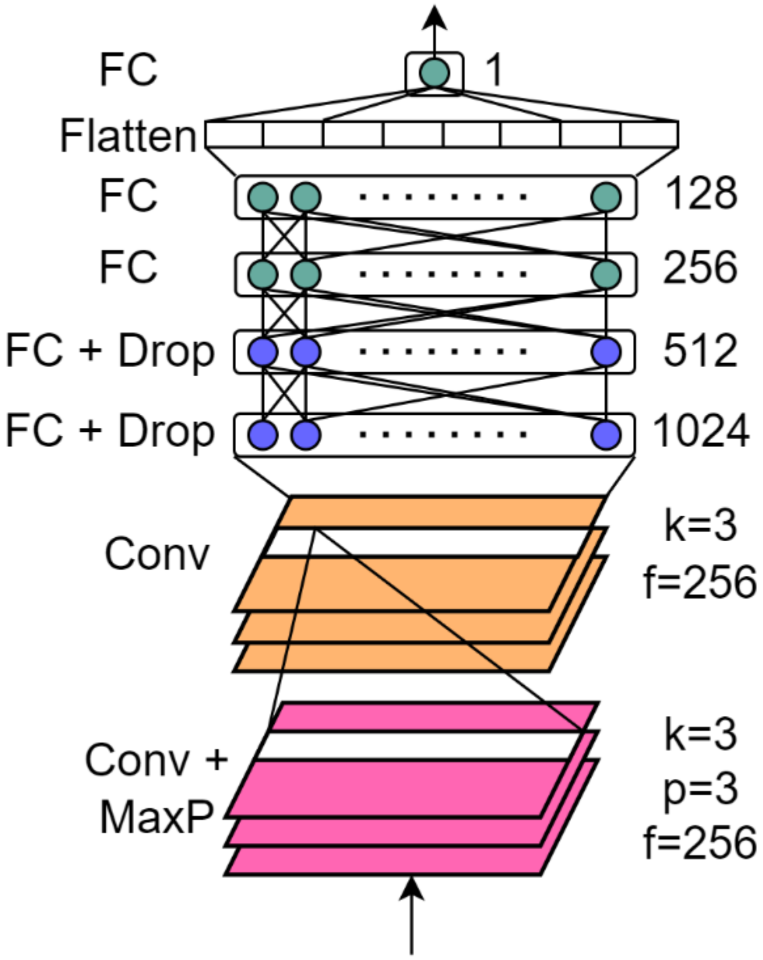


**Supplementary Figure S3: The CNN architecture of ASDSpeech.** Conv: for 1d convolution layer, Conv + MaxP: convolution layer followed by max pooling layer, FC: fully connected layer, FC + Drop: fully connected layer followed by dropout of 0.5, k: kernel size, p: pooling size, f: filters. Two separate models with the same architecture were trained to estimate ADOS-2 SA and ADOS-2 RRB scores respectively.

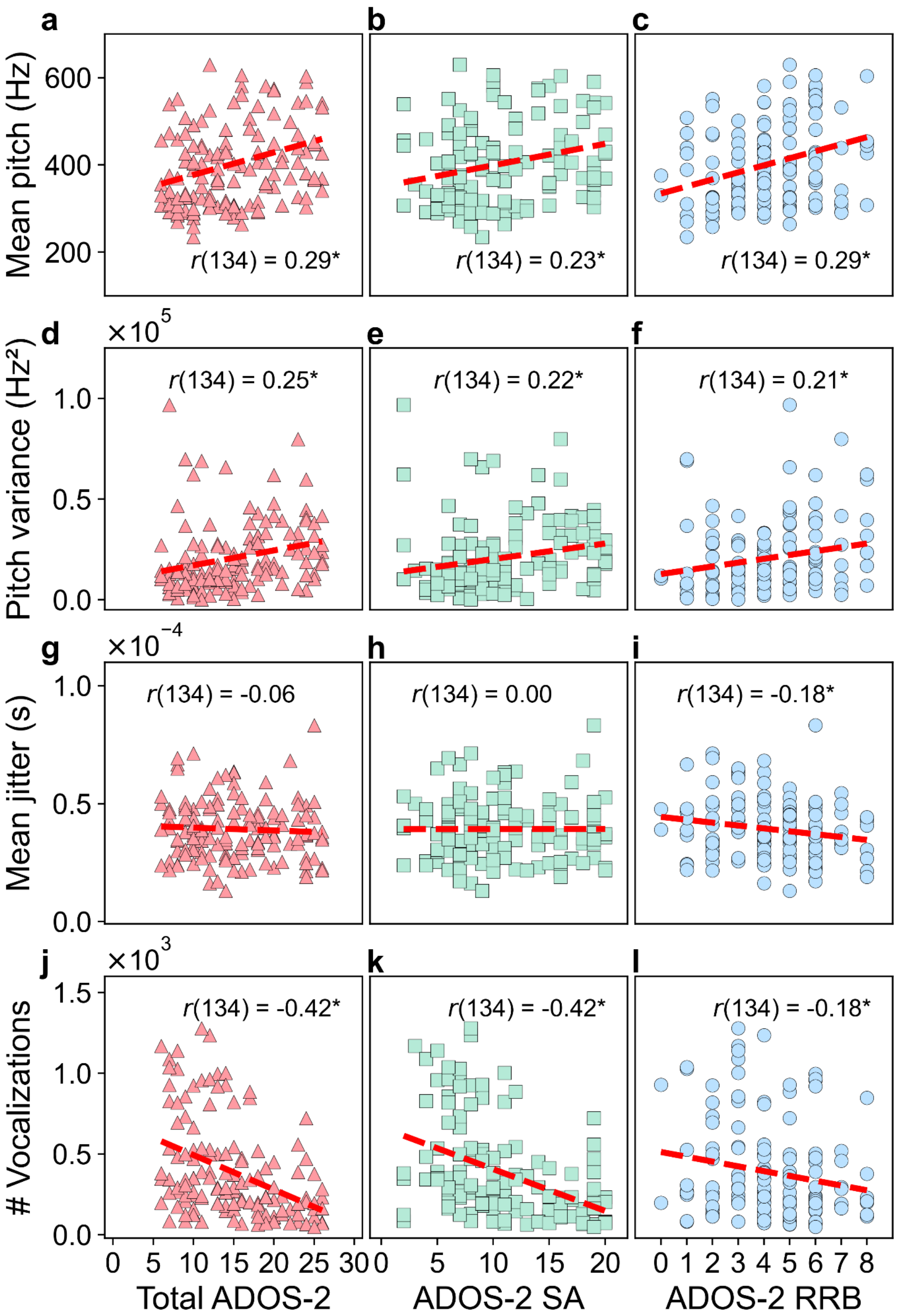


**Supplementary Figure S4: Scatter plots demonstrating the association between each of four different features and total (left column), SA (middle column), or RRB (right column) ADOS-2 scores**. (**a-c**) Mean pitch across voiced vocalizations. (**d-f**) Pitch variance across voiced vocalizations (**g-i**) Mean jitter across all vocalizations. **(k-l)** Number of child vocalizations. Each dot represents a different child/recording from the training set of 136 children who participated in only one ADOS-2 assessment. **Asterisk**: significant Pearson correlation (* < 0.05).

**Supplementary Table S1: Acoustic and conversational features extracted from the child vocalizations.**

| # | Feature type | Feature |
|---|---|---|
| 1 | Pitch (F0) | Mean F0 across consecutive† vocalizations |
| 2 | | Variance of F0 across consecutive vocalizations |
| 3 | | Correlation of coefficient ($\sigma^2/\mu$) across consecutive vocalizations |
| 4 | | Mean of minimum F0: the minimum F0 calculated across vocalization frames and then averaged across consecutive vocalizations |
| 5 | | Variance of minimum F0: the minimum F0 calculated across vocalization frames. Their variance was computed across the minimum F0 values of consecutive vocalizations |
| 6 | | Mean of maximum F0: the maximum F0 calculated across vocalization frames and then averaged across consecutive vocalizations |
| 7 | | Variance of maximum F0: the maximum F0 calculated across vocalization frames. Their variance was computed across the maximum F0 values of consecutive vocalizations |
| 8 | | Mean F0 across VVs within the consecutive vocalizations |
| 9 | | Variance of F0 across VVs within the consecutive vocalizations |
| 10 | | Variance of the mean F0 across VVs: the mean F0 calculated for each vocalization. Their variance was computed across the VVs within the consecutive vocalizations |
| 11 | Formants (F) & Bandwidth (BW) | Mean F1 across consecutive vocalizations |
| 12 | | Variance of F1 across consecutive vocalizations |
| 13 | | Mean F2 across consecutive vocalizations |
| 14 | | Variance of F2 across consecutive vocalizations |
| 15 | | Mean \|F1-F2\| across consecutive vocalizations |
| 16 | | Variance of \|F1-F2\| across consecutive vocalizations |
| 17 | | Mean BW1 across consecutive vocalizations |
| 18 | | Variance of BW1 across consecutive vocalizations |
| 19 | | Mean BW2 across consecutive vocalizations |
| 20 | | Variance of BW2 across consecutive vocalizations |
| 21 | Jitter | Mean jitter across consecutive vocalizations |
| 22 | | Variance of jitter across consecutive vocalizations |
| 23 | Voicing | Mean voicing across consecutive vocalizations |
| 24 | | Variance of voicing across consecutive vocalizations |
| 25 | Energy (E) | Mean E across consecutive vocalizations |
| 26 | | Mean ΔE across consecutive vocalizations |
| 27 | | Mean ΔΔE across consecutive vocalizations |
| 28 | | Mean \|ΔE\| across consecutive vocalizations |
| 29 | | Variance E across consecutive vocalizations |
| 30 | | Variance of ΔE across consecutive vocalizations |
| 31 | | Variance of ΔΔE across consecutive vocalizations |
| 32 | | Variance of \|ΔE\| across consecutive vocalizations |
| 33 | Zero-Crossing Rate (ZCR) | Mean ZCR across consecutive vocalizations |
| 34 | | Variance of ZCR consecutive vocalizations |
| 35 | | Mean ZCR across VVs within consecutive vocalizations |
| 36 | | Variance of ZCR across VVs within the consecutive vocalizations |
| 37 | | Mean ZCR across UVs within the consecutive vocalizations |
| 38 | | Variance of ZCR across UVs within the consecutive vocalizations |
| 39 | Spectral slope (SpSl) | Mean SpSl [range: 20–500 Hz] Hz across consecutive vocalizations |
| 40 | | Variance of SpSl [range: 20–500 Hz] across consecutive vocalizations |
| 41 | | Mean SpSl [range: 50–1500 Hz] across consecutive vocalizations |

| | | |
|---|---|---|
| 42 | | Variance of SpSl [range: 50–1500 Hz] across consecutive vocalizations |
| 43 | | Mean SpSl [range: 20–500 Hz] across VVs within consecutive vocalizations |
| 44 | | Mean SpSl [range: 20–500 Hz]across UVs within consecutive vocalizations |
| 45 | | Mean SpSl [range: 50–1500 Hz] across VVs within consecutive vocalizations |
| 46 | | Mean SpSl [range: 50–1500 Hz] across UVs within consecutive vocalizations |
| 47 | Duration | Mean duration across consecutive vocalizations |
| 48 | | Variance of duration across consecutive vocalizations |
| 49 | Quantity | Total number of vocalizations |

† The number of consecutive vocalizations was set to ten.

VV – Voiced Vocalization, UV – Unvoiced Vocalization.
